# Clinical characteristics of Coronavirus Disease 2019 (COVID-19) patients in Kuwait

**DOI:** 10.1101/2020.06.14.20131045

**Authors:** Abdullah Alshukry, Hamad Ali, Yaseen Ali, Talal Al Taweel, Mohammad Abu-Farha, Jehad AbuBaker, Sriraman Devarajan, Ali A. Dashti, Ali Bandar, Hessah Taleb, Abdullah Al Bader, Nasser Y. Aly, Ebaa Al-Ozairi, Fahd Al-Mulla, Mohammad Bu Abbas

## Abstract

This is a retrospective single-center study of 417 consecutive patients with coronavirus disease 2019 (COVID-19) admitted to Jaber Al-Ahmad Hospital in Kuwait between February 24, 2020 and May 24, 2020. In total, 39.3% of patients were asymptomatic, 41% were symptomatic with mild/moderate symptoms, 5.3% were admitted to the intensive care unit (ICU) and recovered, and 14.4% died. The mean age of death cases was 54.20 years (± 11.09). Comorbidities were more prevalent in patients who died compared with others. Key findings include abnormal levels of markers assicated with infection, inflammation, abnormal blood clotting, heart problems and kidney problems in patients with severe form of the disease and poor putcome. We report a rapidly deteriorating estimated glomerular filtration rate (eGFR) in deaths during ICU stay with kidney injury complications reported in 65% of deaths (*p* < 0.05). Our dynamic profiling of eGFR in ICU highlights the potential role of renal markers in forecasting disease outcome that could perhaps identify patients at risk of poor outcome.

## Introduction

In early December 2019, the first clusters of coronavirus disease 2019 (COVID-19) were identified in Wuhan, China ^1^. These cases were initially reported as pneumonia of unknown cause. They later were attributed to a novel coronavirus, now known as severe acute respiratory syndrome coronavirus 2 (SARS-CoV-2), an enveloped single-strand RNA β-coronavirus with a 30,000-base genome ^2^. On March 11, 2020, the World Health Organization (WHO) confirmed COVID-19 as a pandemic. Since then, the disease has been spreading quickly, affecting more than 5 million people and resulting in more than 350,000 deaths, which emphasizes the threat it poses to global health ^3^.

Clinical manifestations of COVID-19 have shown high variability, including asymptomatic carriers, acute respiratory distress syndrome (ARDS), and pneumonia with variable severity ^4,5^. Most of the identified patients experience mild symptoms, including fever, cough, dyspnea, myalgia, and fatigue. In contrast, patients with severe cases develop ARDS and severe cardiac and renal complications, which can potentially lead to death ^2,6^. Additionally, a poorer prognosis has been associated with older age, being male, and having pre-existing chronic conditions, such as hypertension, cardiovascular disease, and diabetes. At the same time, pediatric cases have shown a milder clinical course ^7^.

Countries worldwide have been affected differently by the COVID-19 pandemic, ranging from high infection and high mortality rates in countries like the USA, France, and Spain to low infection and mortality rates in countries like New Zealand ^3,8^. Multiple theories have been suggested to explain the current infection and death rate, including tighter measures, better health care systems, and genetic factors, among others. One particular hypothesis has focused on genetic variants within the angiotensin converting enzyme 2 (ACE2) gene ^9,10^. Multiple studies have suggested that variants within the ACE2 gene can influence the viral entry into the host cell ^11,12^. In a recent study, we have shown that one variant within the ACE2 gene (N720D) was responsible for reduced infection rates in Middle Eastern countries compared with Europeans ^9^. Such findings highlight the importance of investigating various populations worldwide to gain more insight into this disease.

This study is focused on presenting a cohort from Kuwait, a small country located on the northern tip of the Persian Gulf, with a population of nearly 4.5 million. Kuwait reported its first cases of COVID-19 on February 24, 2020, in passengers coming from Iran. Since then, the reported cases have been increasing exponentially, reaching over 35,000 cases at the time of this report and more than 280 registered deaths ^3,13^. While the search for effective treatment and vaccine for COVID-19 continues, health care systems, including that in Kuwait, are attempting to strengthen their frontline clinical services to cope with the pandemic.

Here, we describe the demographics, baseline comorbidities, clinical characteristics, and outcomes of a cohort of patients with COVID-19 in Kuwait. We further investigate the dynamics of certain laboratory parameters in intensive care unit (ICU) admissions in relation to clinical outcomes.

## RESULTS

### Cohort characteristics

In our study, 417 hospitalized patients with COVID-19 with a positive viral RT-PCR test were enrolled. The cohort had a median age of 47 years (IQR 32–60 years), and 63% of the cohort were men. The age structure of the cohort showed that the majority of patients belonged to the 21– 40 years age group, and notably, only 0.3% belonged to the 81–90 years age group (Fig 1 and Table 1). The cohort consisted of 39.3% asymptomatic patients, 41% symptomatic (mild to moderate), 5.3% admitted to the ICU who recovered, and 14.4% admitted to ICU who died (Table 2).

**Figure 1.**
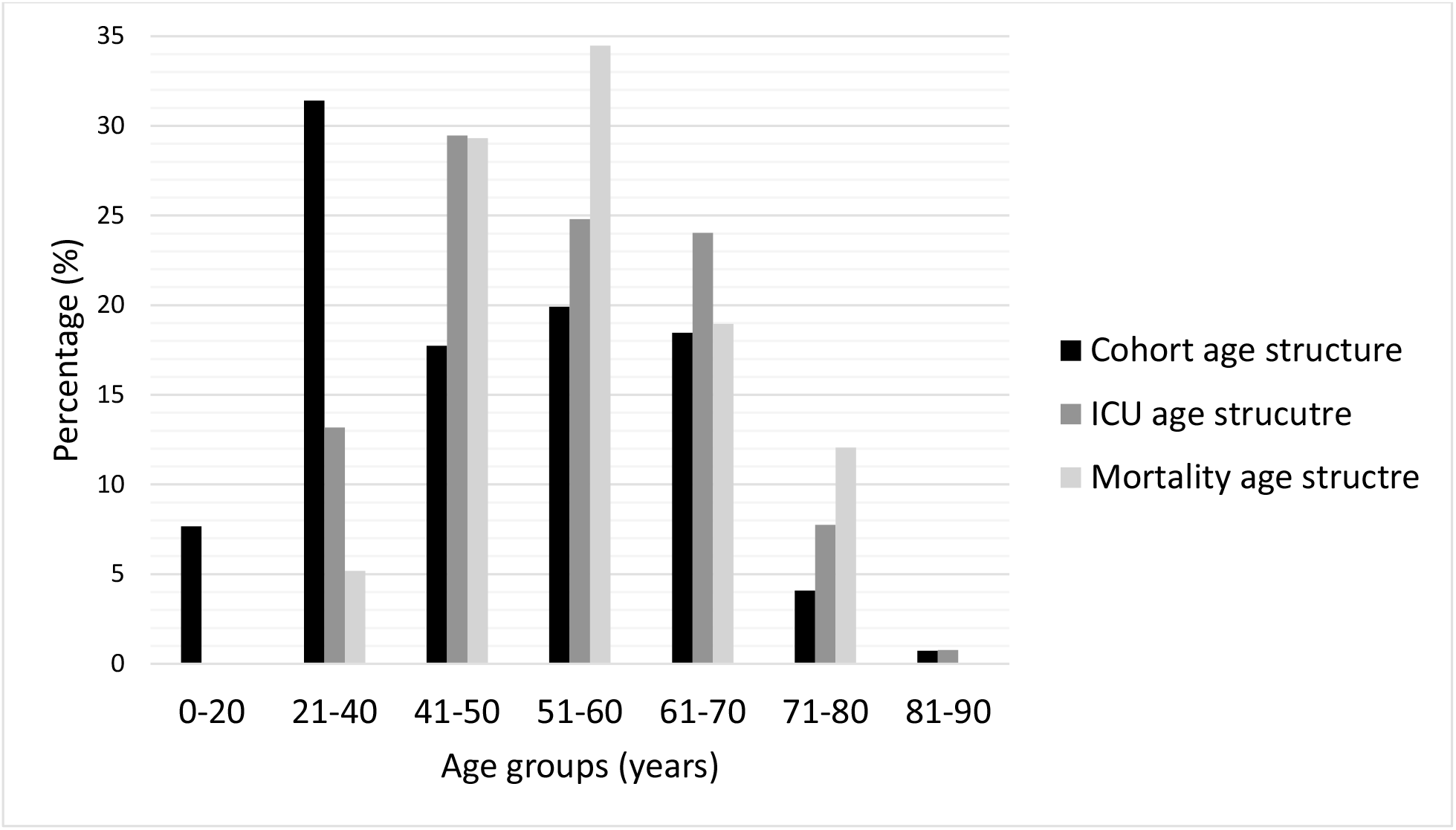
Categorical age structures. Each age group percentage is calculated by dividing the count by the total in each category. Majority of cohort belonged to the age group (21-40 years) while highest numbers of death cases were recorded in the age groups (51-60 years).

**Table 1.** Cohort age structure and Case-fatality rate (CFR) per age group

|  | No. of cases | % of total | No. of ICU admissions | % of total ICU | No. of deaths | % of total deaths | Case-fatality rate, % |
| --- | --- | --- | --- | --- | --- | --- | --- |
| All | 417 | 100 | 129 | 100 | 60 | 100 | 14.4 |
| Age groups, years |  |  |  |  |  |  |  |
| 0-20 | 32 | 7.7 | 0 | 0.0 | 0 | 0.0 | 0.0 |
| 21-40 | 131 | 31.4 | 17 | 13.2 | 5 | 8.3 | 3.8 |
| 41-50 | 74 | 17.7 | 38 | 29.5 | 18 | 30.0 | 24.3 |
| 51-60 | 83 | 19.9 | 32 | 24.8 | 20 | 33.3 | 24.1 |
| 61-70 | 77 | 18.5 | 31 | 24.0 | 11 | 18.3 | 14.3 |
| 71-80 | 17 | 4.1 | 10 | 7.8 | 6 | 10.0 | 35.3 |
| 81-90 | 3 | 0.7 | 1 | 0.8 | 0 | 0.0 | 0.0 |

**Table 2.** Cohort baseline characteristics

|  | Asymptomatic | Symptomatic | ICU Survivors | ICU Death | P Value |
| --- | --- | --- | --- | --- | --- |
| Number N (%) | 164 (39.3) | 171 (41.0) | 22 (5.3) | 60 (14.4) | - |
| Age (Mean ± SD) | 41.97 ± 19.21 | 44.67 ± 15.75 | 52.32 ± 13.51 | 54.20 ± 11.09 | <0.001 |
| **Age Groups** |  |  |  |  |  |
| 0-20 Years | 25 (15.2) | 7 (4.1) | 0 | 0 | - |
| 21 - 40 Years | 56 (34.1) | 65 (38.0) | 5 (22.7) | 5 (8.3) | - |
| 41 - 50 Years | 23 (14.0) | 28 (16.4) | 5 (22.7) | 18 (30.0) | - |
| 51 - 60 Years | 24 (14.6) | 35 (20.5) | 4 (18.2) | 20 (33.3) | - |
| 61 - 70 Years | 27 (16.5) | 32 (18.7) | 7 (31.8) | 11 (18.3) | - |
| 71 - 80 Years | 7 (4.3) | 3 (1.8) | 1 (4.5) | 6 (10.0) | - |
| 81+ Years | 2 (1.2) | 1 (0.6) | 0 | 0 | - |
| **Gender** |  |  |  |  |  |
| Male | 86 (52.4) | 107 (62.6) | 15 (68.2) | 54 (90.0) | - |
| Female | 78 (47.6) | 64 (37.4) | 7 (31.8) | 6 (10.0) | - |
| **Signs & Symptoms** |  |  |  |  |  |
| Fever | - | 94 (55.0) | 12 (54.5) | 37 (61.7) | 0.652 |
| Chills | - | 7 (4.1) | 0 (0.0) | 4 (6.7) | 0.483 |
| Dysgeusia | - | 1 (0.6) | 0 (0.0) | 0 (0.0) | 0.684 |
| Dry Cough | - | 89 (52.0) | 12 (54.5) | 35 (58.3) | 0.699 |
| Productive Cough | - | 21 (12.4) | 2 (9.1) | 3 (5.0) | 0.226 |
| Headache | - | 24 (14.1) | 0 (0.0) | 3 (5.0) | 0.065 |
| Chest Pain | - | 5 (2.9) | 0 (0.0) | 5 (8.3) | 0.132 |
| Syncope | - | 0 (0.0) | 0 (0.0) | 1 (1.7) | 0.237 |
| Runny Nose | - | 13 (7.6) | 0 (0.0) | 1 (1.7) | 0.122 |
| Sore Throat | - | 51 (29.8) | 0 (0.0) | 10 (16.7) | 0.060 |
| Shortness of Breadth | - | 14 (8.2) | 13 (59.1) | 49 (81.7) | <0.001 |
| Nausea | - | 6 (3.5) | 3 (13.6) | 3 (5.0) | 0.197 |
| Vomiting | - | 4 (2.3) | 2 (9.1) | 4 (6.7) | 0.176 |
| Abdominal Pain | - | 6 (3.5) | 2 (9.1) | 4 (6.7) | 0.411 |
| Diarrhea | - | 9 (5.3) | 2 (9.1) | 2 (3.3) | 0.594 |
| Fatigue | - | 20 (11.7) | 4 (18.2) | 16 (26.7) | 0.030 |
| Myalgia | - | 37 (21.6) | 2 (9.1) | 9 (15.0) | 0.215 |
| **Hospital Admission** |  |  |  |  |  |
| Days (Mean ± SD) | 20.69 ± 8.57 | 21.40 ± 8.28 | 11.95 ± 8.96 | 13.15 ± 10.02 | <0.001 |
| **Comorbidities** |  |  |  |  |  |
| Diabetes | 36 (22.2) | 32 (18.8) | 5 (22.7) | 24 (40.0) | 0.016 |
| Hypertension | 40 (24.5) | 50 (29.2) | 5 (22.7) | 28 (46.7) | 0.016 |
| Asthma | 14 (18.5) | 12 (7.1) | 3 (13.6) | 12 (20.3) | 0.046 |
| COPD | 0 | 0 | 0 | 1 (1.7) | 0.271 |
| Cardiovascular disease | 7 (4.3) | 18 (10.6) | 1 (4.5) | 13 (21.7) | 0.002 |
| Chronic renal disease | 1 (0.6) | 8 (4.7) | 1 (4.5) | 4 (6.7) | 0.041 |
| Malignancy | 4 (2.4) | 4 (2.4) | 1 (4.5) | 3 (5.0) | 0.734 |

The majority (29.5%) of ICU admissions belonged to the 41–50-year age group, whereas 24.8% were 51–60 years and 24% were 61–70 years of age (Fig 1 and Table 1). No ICU admissions were recorded for the youngest age group (0–20 years).

Of 417 cases, we recorded 60 deaths, the majority of such patients belonging to the age group 51–60 years, which represented 33.3% of all deaths reported, with a case-fatality rate of 24.1% (Fig 1 and Table 1). However, the case-fatality rate was highest in those aged 71–80 years, at 35.3% (Table 1).

The patients with COVID-19 were divided into four groups. The first group was composed of asymptomatic patients. These patients had a positive viral RT-PCR but did not show any symptoms and were kept in hospital isolation until a negative viral RT-PCR was achieved, with an average hospital admission length of 20.69 ± 8.57 days (Table 2). The asymptomatic group made up 39.3 % of the cohort, with an average age of 41.97 ± 19.21 years. Symptomatic cases made up 41% of the cohort. This group consisted of patients who exhibited mild/moderate symptoms but did not require ICU admission. Some 55% of these patients experienced fever, 52% presented with a dry cough, 29.8% reported a sore throat, and 21.6% described symptoms of myalgia (Table 2). Cases admitted to the ICU were divided into two groups based on outcome: the ICU survivors (composed of 22 patients, 5.3% of the cohort) and the ICU deaths (composed of 60 patients, accounting for 14.4% of the cohort) (Table 2). The pattern of symptoms in the ICU survivors’ group was similar to the symptomatic group except for the shortness of breath, which was present in 59.1% of cases (Table 2). Shortness of breath was more prevalent in the ICU deaths group (81.7%). Furthermore, this group had more comorbidities than the other groups, including diabetes, hypertension, asthma, cardiovascular disease, chronic renal disease, and malignancies (Table 2).

### Clinical findings

The clinical data analysis indicated significant differences between the COVID-19 groups. The asymptomatic group generally showed normal laboratory findings with minimal borderline abnormalities (Table 3). Numerous markers showed significant differences between the symptomatic/mild group and patients admitted to the ICU, including complete blood count (Table 3). Inflammatory markers, including procalcitonin (PCT) and C-reactive protein (CRP) showed progressive increasing patterns from the symptomatic to the ICU deaths group. Markers of blood clotting, including prothrombin time (PT) and activated partial thromboplastin time (APTT) showed prolonged timings in the group of ICU deaths.

**Table 3.** Laboratory findings of COVID-19 patients groups

|  | Reference range /unit | Asymptomatic |  | Symptomatic |  | ICU Survivor |  | ICU Death |  | p value |
| --- | --- | --- | --- | --- | --- | --- | --- | --- | --- | --- |
|  |  | n | Median (IQR) | n | Median (IQR) | n | Median (IQR) | n | Median (IQR) |  |
| White blood cells (WBC) | 3.7-10 x10<sup>9</sup> /L | 164 | 6.24 (5.26 - 7.64) | 171 | 6.2 (5.3 - 7.4) | 22 | 8.08 (6.45 - 9) | 60 | 12.68 (9.45 - 16.05) | <0.001 |
| Neutrophils | 1.7 – 6 x10<sup>9</sup> /L | 164 | 2.92 (2.30 - 3.38) | 171 | 3.36 (2.64 - 4.1) | 22 | 5.37 (4.38 - 6.64) | 60 | 10.96 (8.02 - 14.21) | <0.001 |
| Lymphocytes | 1 -3 x10<sup>9</sup> /L | 164 | 2.39 (1.89 - 2.89) | 171 | 2.05 (1.74 - 2.5) | 22 | 1.41 (1.14 - 1.93) | 60 | 0.85 (0.68 - 1.05) | <0.001 |
| Monocytes | 0.2 – 1 x10<sup>9</sup> /L | 164 | 0.57 (0.48 - 0.7) | 171 | 0.55 (0.46 - 0.7) | 22 | 0.57 (0.5 - 0.79) | 60 | 0.57 (0.35 - 0.79) | 0.835 |
| Eosinophils | 0.02 – 0.5 x10<sup>9</sup> /L | 164 | 0.15 (0.1 - 0.24) | 171 | 0.12 (0.08 - 0.19) | 22 | 0.13 (0.09 - 0.22) | 60 | 0.05 (0.01 - 0.1) | <0.001 |
| Basophils | 0.5% to 1% | 164 | 0.45 (0.35 - 0.58) | 171 | 0.4 (0.3 - 0.56) | 22 | 0.33 (0.25 - 0.54) | 60 | 0.22 (0.17 - 0.3) | <0.001 |
| Platelets | 130-430 x10<sup>9</sup> /L | 164 | 267.27 (225.04 - 315.52) | 171 | 274 (224.8 - 331.33) | 22 | 319.28 (269.03 - 388.08) | 60 | 260.51 (204.03 - 316.34) | 0.0213 |
| Red blood cells (RBC) | 4.5-5.5 x10<sup>12</sup> /L | 164 | 5.03 (4.52 - 5.43) | 171 | 5.04 (4.6 - 5.28) | 22 | 4.48 (4.19 - 4.82) | 60 | 3.85 (3.36 - 4.35) | <0.001 |
| Hemoglobin (HB) | 130-170 g/L | 164 | 133.83 (120.54 - 145.51) | 171 | 134.17 (123.33 - 145.5) | 22 | 119.03 (105.96 - 128.63) | 60 | 104.93 (87.08 - 120.39) | <0.001 |
| Prothrombin Time (PT) | 10-13 s | 49 | 13.4 (12.7 - 14.1) | 44 | 13.35 (12.56 - 14.75) | 17 | 13.63 (12.8 - 14.5) | 54 | 15.23 (13.95 - 19.05) | <0.001 |
| Activated partial thromboplastin time (APTT) | 23-30 s | 49 | 32 (30.5 - 35.55) | 45 | 32.5 (29.4 - 38.68) | 17 | 32.05 (30 - 36.41) | 54 | 45.2 (38.94 - 53.91) | <0.001 |
| D-dimer | 0-500 mg/L | 11 | 186 (155 - 436) | 30 | 331.5 (212.5 - 483.38) | 11 | 682 (555 - 1668) | 40 | 1987 (817.25 - 5037.33) | <0.001 |
| C-reactive protein (CRP) | <10 mg/L | 162 | 3.5 (2 - 8.69) | 167 | 9.88 (4 - 38) | 22 | 88.22 (45.52 - 106.62) | 44 | 211.33 (133.83 - 303.03) | <0.001 |
| Procalcitonin (PCT) | <0.05 ng/ml | 154 | 0.04 (0.03 - 0.06) | 160 | 0.06 (0.03 - 0.09) | 22 | 0.27 (0.08 - 0.46) | 60 | 3.36 (1.13 - 9.61) | <0.001 |
| Urea | 2.5-7.1 mmol/L | 164 | 3.33 (2.83 - 4.03) | 171 | 3.56 (2.95 - 4.24) | 22 | 4.9 (4.31 - 7) | 60 | 15.56 (9.56 - 22.97) | <0.001 |
| eGFR | >90 mL/min/1.73m<sup>2</sup> | 147 | 105.8 (95.5 - 117.25) | 165 | 104.43 (92.78 - 114.25) | 22 | 94.54 (67.35 - 104.58) | 60 | 47.63 (26.26 - 81.19) | <0.001 |
| Creatinine | 74.3-107 mmol/L | 164 | 61.46 (51.5 - 75.86) | 171 | 70.5 (58.38 - 78.5) | 22 | 78.83 (69.44 - 86.9) | 60 | 193.63 (99.02 - 317.43) | <0.001 |
| Total Protein | 60-80 g/L | 164 | 65.33<br>(62.08 -<br>68.15) | 171 | 64.67<br>(61.67 -<br>68.33) | 22 | 61.6<br>(57.45 -<br>63.26) | 60 | 56.36<br>(51.54 -<br>59.37) | <0.001 |
| Albumin | 35-55 g/L | 164 | 37.52<br>(35.12 -<br>40.3) | 171 | 35.52<br>(32.1 -<br>38.26) | 22 | 27.28<br>(26.6 -<br>30.65) | 60 | 20.09<br>(18.38 -<br>23.13) | <0.001 |
| Alkaline<br>phosphatase<br>(ALP) | 20-140 IU/L | 164 | 62.17<br>(51.19 -<br>73.48) | 171 | 57.25<br>(48.86 -<br>73) | 22 | 72.51<br>(51.38 -<br>80.93) | 60 | 96.4<br>(74.07 -<br>135.91) | <0.001 |
| Alanine<br>aminotransferase<br>(ALT) | 7-56 IU/L | 164 | 20.71<br>(15.08 -<br>28.5) | 171 | 27.33<br>(19.33 -<br>42.71) | 22 | 39.46<br>(31.47 -<br>62.58) | 60 | 41.99<br>(29.67 -<br>84.35) | <0.001 |
| Aspartate<br>aminotransferase<br>(AST) | 10-40 IU/L | 164 | 21.45<br>(18.25 -<br>25.78) | 171 | 24.5<br>(20.33 -<br>33) | 22 | 36.4<br>(27.38 -<br>47) | 60 | 56.75<br>(45.47 -<br>104.71) | <0.001 |
| Gamma-glutamyl<br>transferase (GGT) | 9-48 IU/L | 164 | 15.83<br>(11.46 -<br>21.89) | 170 | 23.29<br>(14.92 -<br>39.86) | 22 | 45.05<br>(32.75 -<br>94.77) | 60 | 76.5<br>(45.3 -<br>115.88) | <0.001 |
| Total Bilirubin | 5-21 mmol/L | 164 | 11.59<br>(9.43 -<br>14.63) | 171 | 11.7 (9.32<br>- 14.01) | 22 | 14.86<br>(12.36 -<br>17.25) | 60 | 21.18<br>(14.79 -<br>33.91) | <0.001 |
| Direct Bilirubin | 1.7-5.1 µmol/L | 164 | 1.75 (1.25<br>- 2.4) | 171 | 2.03 (1.38<br>- 2.7) | 22 | 4.2<br>(3.25 -<br>5.84) | 60 | 9.14<br>(5.88 -<br>18.88) | <0.001 |
| TROPONIN | 0-0.4 ng/mL | 5 | 0.01 (0.01<br>- 4) | 29 | 2.67 (0.01<br>- 5.42) | 10 | 5 (0.01<br>- 6.97) | 25 | 56 (22 -<br>164.45) | <0.001 |
| Lactate<br>dehydrogenase<br>(LDH) | 140-280 IU/L | 36 | 172<br>(144.61 -<br>207.61) | 48 | 214.43<br>(166.75 -<br>277) | 12 | 325<br>(293.96 -<br>478.88) | 27 | 547<br>(412.79 -<br>633.33) | <0.001 |

Markers related to renal function showed significant abnormalities in the ICU deaths, including a reduced estimated glomerular filtration rate (eGFR) and increased urea (Table 3), which coincide with reported kidney injury complications in 65% of cases in this particular group (Table 4). Markers associated with heart injury, including troponin and lactate dehydrogenase, showed significant increases in patients admitted to the ICU and in particular, the ICU deaths (Table 3), which coincided with reported heart failure in 86% of cases in this specific group and cardiac injury in 44.1% of them (Table 4). D-dimer was within the normal range in the asymptomatic and symptomatic groups but was significantly elevated in the ICU group and the ICU deaths (Table 3).

**Table 4.** Complications and treatments in COVID-19 patients’ groups

|  | Asymptomatic | Symptomatic | ICU Survivor | ICU Death | P value |
| --- | --- | --- | --- | --- | --- |
|  | Number (%) | Number (%) | Number (%) | Number (%) |  |
|  | 164 (39.3) | 171 (41.0) | 22 (5.3) | 60 (14.4) |  |
| Complications |  |  |  |  |  |
| Sepsis | NA | NA | 10 (45.5) | 46 (79.3) | <0.001 |
| ARDS | NA | NA | 18 (81.8) | 60 (100.0) | <0.001 |
| Heart Failure | NA | NA | 3 (13.6) | 49 (86.0) | <0.001 |
| Cardiac Injury | NA | NA | 2 (9.1) | 26 (44.1) | <0.001 |
| Kidney Injury | NA | 1 (0.6) | 1 (4.5) | 39 (65.0) | <0.001 |
| Treatment |  |  |  |  |  |
| Antibiotic | 10 (6.1) | 54 (31.6) | 19 (95.0) | 51 (94.4) | <0.001 |
| Antiviral | 7 (4.2) | 37 (21.6) | 18 (90.0) | 38 (70.4) | <0.001 |
| Antimalarial | 3 (1.8) | 24 (14.0) | 12 (60.0) | 37 (68.5) | <0.001 |
| Corticosteroids | NA | 2 (1.2) | 3 (15.0) | 1 (1.9) | 0.003 |
| Plasma | NA | NA | NA | 2 (3.8) | 0.125 |
| ECMO | NA | NA | 1 (5.0) | 2 (3.7) | 0.015 |
| Intubation (Yes) | NA | NA | 11 (55.0) | 51 (94.4) | <0.001 |
| Tracheostomy (Yes) | NA | NA | 1 (5.0) | 6 (11.1) | <0.001 |

A total of 81.8% of patients in the ICU survivor group developed ARDS compared with 100% of cases in the ICU deaths (Table 3). Chest radiographs of patients in the ICU survivor and death groups showed diffuse bilateral airspace opacification with patches of consolidation (Fig 2). High-resolution chest computed tomography in the ICU deaths group showed multifocal large patchy areas of ground glass opacity mixed with dense consolidations (Fig 2). Intubation was required for 55% of the patients in ICU survivor group and in 94.4% of those in the ICU deaths group (Table 3). Treatment-wise, antibiotics such as amoxicillin, Augmentin, Rocephin, and piperacillin/tazobactam were the most commonly administered medications, with 31.6% of the symptomatic, 95% of the ICU survivors, and 94.4% of the ICU deaths group receiving them. Antiviral drugs, such as oseltamivir and lopinavir, were administered to 90% of patients in the ICU survivor group and in 70.4% of the ICU deaths group. The antimalarial drug hydroxychloroquine was administered to 60% of the ICU survivor group and 68.5% of the ICU deaths group (Table 4).

**Figure 2.**
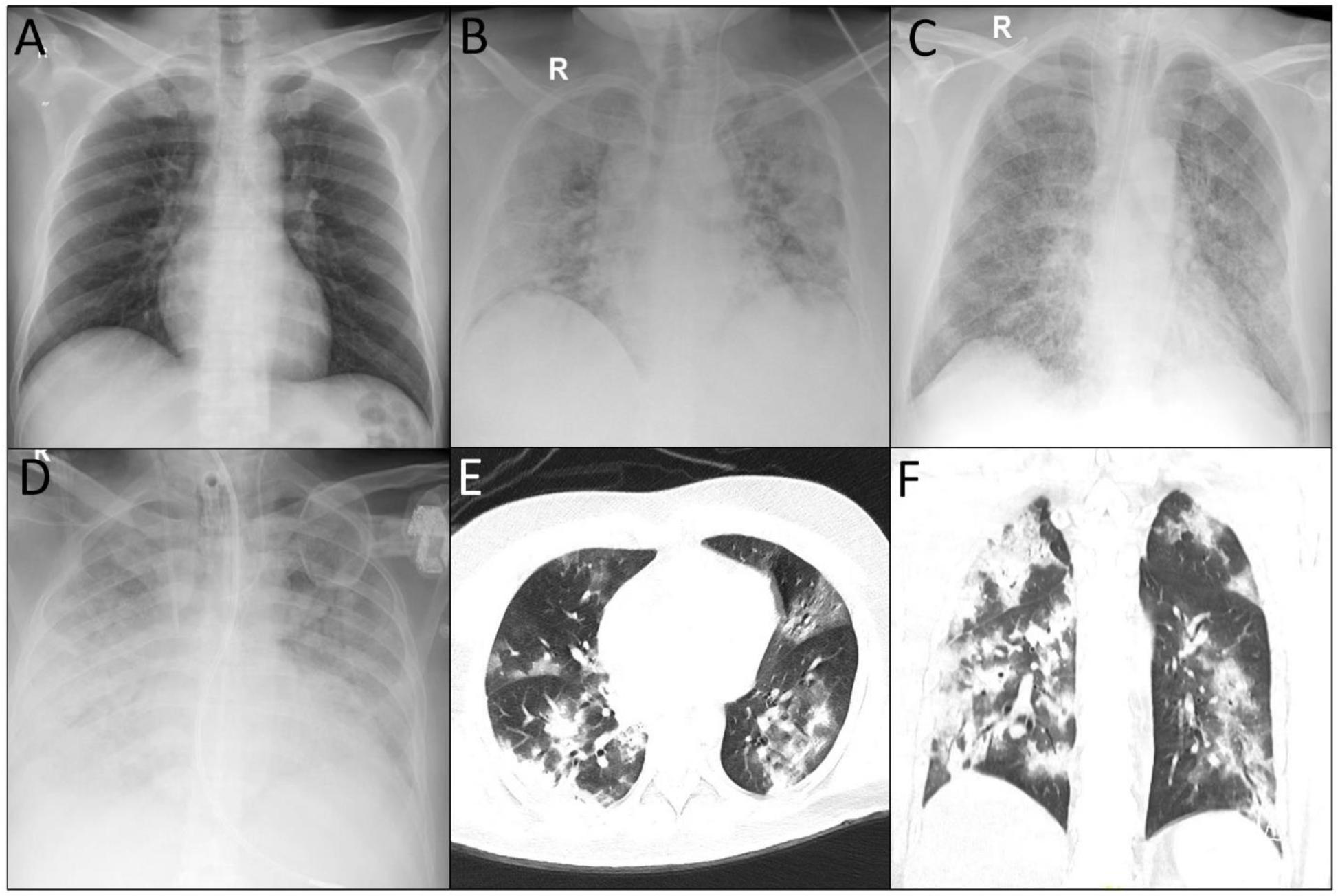
COVID-19 patients chest imaging. A. Normal chest radiography of a patient with mild to moderate COVID-19. B. Diffuse bilateral airspace opacification with patches of consolidation in an ICU survivor. C. Diffuse bilateral airspace opacification with consolidation patches in an ICU death. D. Diffuse bilateral airspace opacification with near total white out of both lung fields in an ICU death. E and F. High Resolution CT chest showing multifocal large patchy areas of ground glass opacification mixed with dense consolidations in an ICU death.

### Dynamic laboratory profiles of ICU admissions

We tracked the levels of six blood markers associated with infection, inflammation, and kidney function in ICU patients to study disease progression and outcome. Patients in the ICU deaths group showed elevated levels of white blood cells after the second day of disease onset onward. In contrast, the ICU survivor group had an average below 10 × 10^9^ cells/L (Fig 3A). A similar pattern was observed in the neutrophil count (Fig 3B). The ICU deaths group also showed lymphocytopenia compared with ICU survivors (Fig 3C). Inflammatory markers, namely CRP and PCT, showed a significant increase during ICU stay until outcome in the ICU deaths group (Fig 3D and 3E). Renal function declined progressively in the ICU deaths group from admission until outcome (Fig 3F).

**Figure 3.**
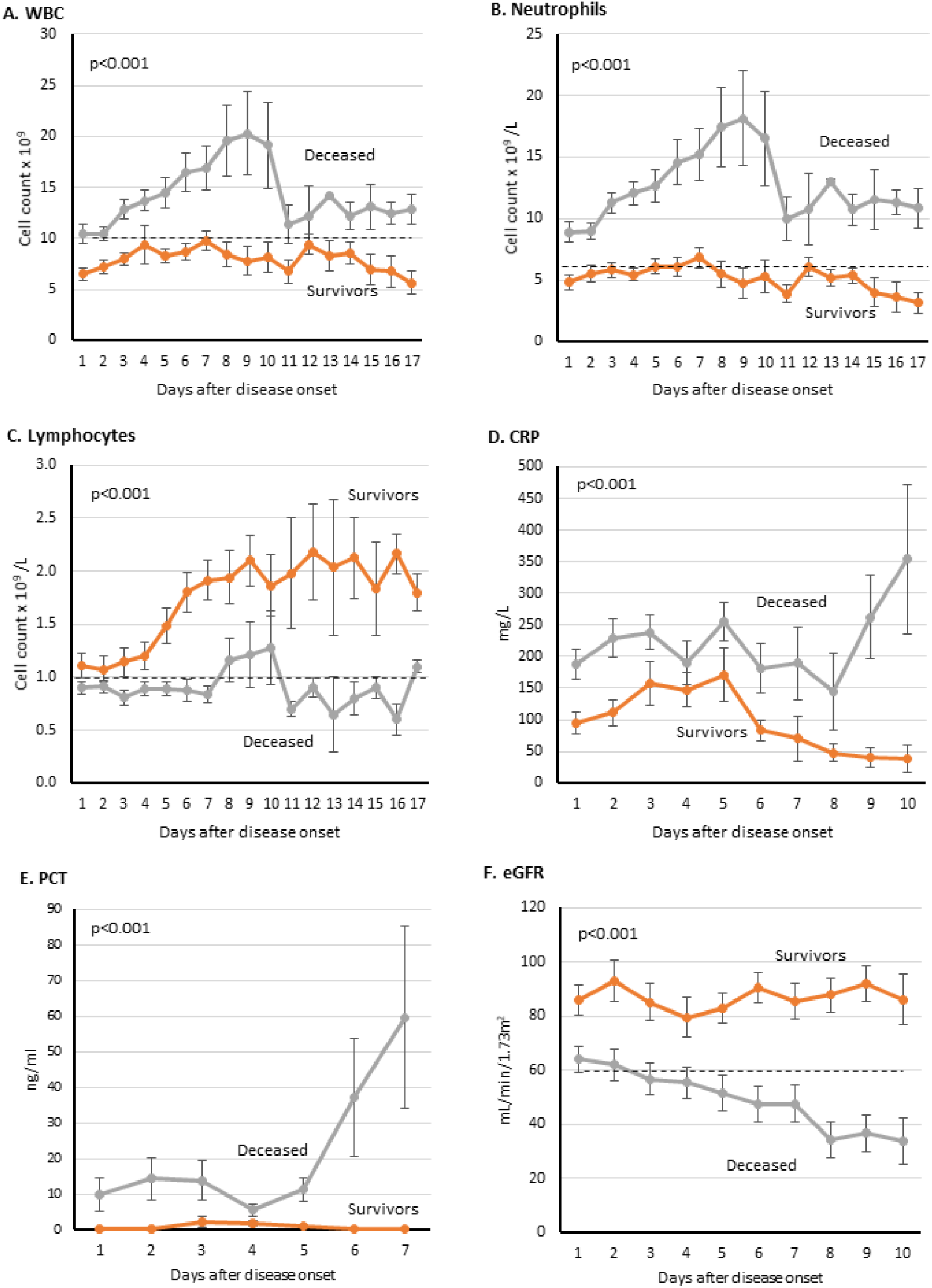
Dynamic Profile of certain laboratory markers in COVID-19 ICU patients. Laboratory tests were done on daily basis after disease onset. Dotted black lines indicate upper normal limit in A and B, and lower normal limit in C and F.

## DISCUSSION

To our knowledge, this study is one of the first detailed reports of COVID-19 clinical characteristics in Kuwait and the region. Kuwait reported its first cases from travelers arriving from Iran on February 24, 2020, followed by further cases arriving from Europe before cases from community transmission began to accumulate ^13^. We recruited our cohort between February 24, 2020 and May 24, 2020, during which the hospitalization rate was 100% for all patients positive for SARS-CoV-2. Detecting all the cases during a specific period and from one place allows more comprehensive clinical insights, especially in relation to the inclusion of asymptomatic cases, which were isolated in the hospital with full medical surveillance.

The most prevalent symptoms in our cohort were fever, which was present in 34.3% of the cohort, dry cough (32.6%), and shortness of breath (18.2%), with the latter being more prevalent in ICU admissions (Table 2). Prevalence of symptoms varied between studies; for instance, fever was reported in 43.8% of patients on admission in a multicenter study in China ^17^, whereas another study conducted in the city of Wuhan found a prevalence of 98.6% ^6^. These variable findings are influenced by the diagnostic criteria used and the effectiveness of surveillance strategies. It is more a question of which led to admission: a symptom or random surveillance testing based on viral PCR resulting in the detection of asymptomatic individuals.

The age structure of the cohort provided a good representation of the population in Kuwait (Fig 1 and Table 1) ^18^. The asymptomatic group represented 39.3% of the reported cases, which is close to the 42.3% reported in Wuhan, the 40% reported in the USA, and the 41% reported in Italy ^19,20^. Detecting asymptomatic individuals is challenging, given these people do not raise any flags for medical attention; therefore, they can intensify infection ^21^. In Kuwait, extensive viral RT-PCR testing was conducted for every passenger entering the country during the period of recruitment, in addition to active contact tracing being enacted.

The case-fatality rate calculated for the cohort was 14.4%, which could represent an overestimation due to the proportion of ongoing cases, especially at the early stage of the outbreak ^22^. The highest case-fatality rate was recorded in the 70–80-year age group (35.3%), which is close to what has been reported in New York City ^23^.

ARDS was one of the main causes for ICU admissions, given it was reported in all deaths and in 81.8% of ICU survivors. Patients admitted to the ICU were older in comparison to other groups. Our results also indicated that the ICU deaths group had a higher prevalence of comorbidities and were predominantly male (Table 2). Several studies have shown a significant association between poorer outcome and being male. This was the case in 2003 during the SARS coronavirus outbreak and now also with COVID-19 ^24^. Although the reasons underlying this observation are not fully understood, the ACE2 level in males has been suggested as a possible explanation ^24^. A death outcome also correlated with higher prevalence of comorbidities, as observed in the ICU deaths group, with hypertension being the most prevalent comorbidity, followed by diabetes. Similar findings have also been reported in New York ^23^ and Wuhan ^6^.

Direct comparison between groups revealed than ICU admissions and particularly the ICU death group indicated significantly lower levels of hemoglobin (Table 3). This observation has been reported previously in patients with other types of pneumonia, associated with higher mortality rate ^25^. The ICU deaths group also showed prolonged PT and activated partial thromboplastin time (APPT) compared with other groups (Table 3), which indicate abnormalities in coagulation. Such abnormal coagulation parameters have been reported by previous studies, which showed an association with poor prognosis ^26^. Recent recommendations advise that such prolonged APPT should not impede the use of anticoagulation therapies to prevent or treat venous thrombosis in patients with COVID-19 ^27^. D-dimer levels were also significantly elevated in the ICU deaths group (Table 3). Previous studies have indicated that patients who required intubation and have higher levels of D-dimer would have a higher risk of developing pulmonary embolism ^28^, which highlights the need for a protocol to identify and treat such cases with anticoagulants.

For ICU admissions, a dynamic analysis of a set of markers was performed throughout the ICU stay until an outcome was reached (recovery or death). In the ICU deaths group, the WBC and neutrophil counts continued to increase until day 10, when the averages dropped but remained above the upper normal limit, which with the decreasing lymphocytes indicate active ongoing infection. PCT levels continued to increase in the ICU deaths group until an outcome was concluded, which could be caused by bacterial coinfection and has been associated with severe disease outcome^29^. CRP also followed a similar increasing pattern, which could indicate a severe inflammatory cascade possibly associated with ARDS (Table 4) and eventually leading to death ^30^.

Kidney injury was more prevalent in the ICU deaths group compared with the ICU survivor and symptomatic groups (65% vs. 4.5% and 0.6%, respectively). This matched the reported progressive impairment of kidney function in the ICU deaths group, as suggested by the declining eGFR low total protein and albumin and increased urea (Fig 3, Tables 3 and 4). ACE2 might play an important role in kidney involvement in COVID-19, particularly in severe cases, given it is highly expressed in the tubular epithelial cells of the kidneys ^31^. When SARS-CoV-2 infects the renal epithelial cells through the ACE2 receptor, it can initiate a cascade of pathological events, including mitochondrial impairment, tubular necrosis, collapsing glomerulopathy, and eventually kidney injury and impairment ^32^. Remarkably, our reported kidney injury prevalence in the ICU group exceeded what has been reported by studies in the USA and Europe, where the prevalence ranged from 20% to 40% of patients admitted to the ICU ^23,32^. This could be attributed to the high prevalence of cardiovascular disease in Kuwait, as suggested by the 41% proportional mortality ^33^. Moreover, 46.7% of the patients in the ICU deaths group had a history of hypertension, which could negatively impact the deteriorating kidney function ^34^. The burden of kidney injury on COVID-19 prognosis should not be underestimated, given our results indicated that kidney injury is a negative prognostic factor for survival. Therefore, therapeutic and preventive protocols need to be developed and adapted to control the burden of kidney injury and to reduce morbidity and mortality.

The pattern of reported markers could highlight possible pathological mechanisms leading to death in patients with COVID-19. Thus far, there is no effective treatment for COVID-19. Patients are provided with supportive care, and efforts are directed toward controlling the infection by antibiotics, antivirals, and antimalarial drugs without noticeable favorable outcome.

## Conclusion

Here, we provide a detailed clinical analysis of a cohort of 417 consecutively recruited patients with COVID-19 from a single hospital in Kuwait. In total, 39.3% of patients were asymptomatic, 41% were symptomatic with mild symptoms, 5.3% were admitted to the ICU and recovered, and 14.4% died. Notably, kidney damage was the most prevalent complication reported in the patients who died, which is supported by renal marker laboratory findings. Our dynamic profiling of eGFR in ICU patients with COVID-19 highlights the potential role of renal markers in forecasting disease outcome and perhaps identifying patients at risk of poor outcome.

## METHODS

### Study design

This was a retrospective study, reviewed and approved by the standing committee for coordination of health and medical research at the Ministry of Health in Kuwait (IRB 2020/1404). The requirement of written informed consent was waived by the standing committee for coordination of health and medical research at the Ministry of Health in Kuwait due to the urgency of data collection and nature of disease being studied. All methods involving the human participants was carried out in accordance with the relevant guidelines and regulations. To our knowledge our cohort data have not been used in any previous reports. The medical records of all confirmed COVID-19 cases admitted to Jaber Al-Ahmad Hospital in Kuwait between February 24 and May 24, 2020 were included in the study. The COVID-19 diagnoses were established based on a positive result of real-time reverse transcriptase-polymerase chain reaction (RT-PCR) assay of nasal and/or pharyngeal swabs, in accordance with WHO interim guidance ^14^. Cases were divided into four groups: asymptomatic, symptomatic with mild/moderate symptombs, ICU survivors and ICU death.

### Data collection

A total of 417 patients with confirmed COVID-19 were included in the study. Patients’ medical records were accessed and analyzed by the research team at Dasman Diabetes Institute, Kuwait University and at Jaber Al-Ahmad Hospital. Epidemiological, clinical, laboratory, and radiological characteristics, in addition to treatment plans and outcomes, were accessed and obtained from the medical records.

Recorded information included demographic data, medical history including underlying comorbidities; travel history; contact tracing data; clinical chemistry; hematological laboratory findings; chest radiological images; treatments; complications; and ICU admissions, durations, and dynamics of hospital stay and outcomes. Signs, symptoms, and laboratory findings were recorded on the day of hospital admission (ward and ICU). ARDS was determined in accordance with the Berlin definition ^15^. Acute kidney injury was determined in accordance with the Kidney Disease: Improving Global Outcomes definition ^16^. Cardiac injury was concluded based on blood cardiac markers, electrocardiography, and/or echocardiography ^6^.

### Hospitalization dynamics

During the period in which patients were enrolled in this study, a 100% hospitalization policy was implemented at Jaber Al-Ahmad Hospital by the Ministry of Health. Any patient with a positive RT-PCR test was admitted, isolated, and put under clinical surveillance, including asymptomatic cases. Patients with mild to moderate COVID-19 symptoms who were hemodynamically stable and without any signs of respiratory distress were admitted to the ward after RT-PCR confirmation for isolation and clinical surveillance and re-evaluation. Patients in this subgroup were transferred to the ICU only if they developed signs of respiratory distress and desaturation of oxygen levels (confirmed by pulse oximetry and arterial blood gases) and/or signs of hemodynamic instability requiring close monitoring and intensive re-establishment of homeostasis. Patients with severe to critical COVID-19 symptoms were directly admitted to the ICU should they meet any of the following criteria of severity: hypoxemic respiratory failure that required respiratory support, such as patients who developed ARDS; hemodynamic instability due to cardiogenic or septic shock and clinical/radiological/laboratory evidence of heart failure; acute cardiac injury; and acute kidney injury secondary to COVID-19 manifestations. For ICU dynamics analysis, we divided patients into two groups: ICU survivors and ICU deaths. Clinical analysis of blood samples was performed for all patients on a daily basis. The daily values of selected laboratory parameters were averaged and plotted until an outcome was achieved.

### Statistical analysis

The variables analysed in the study were divided into categorical and continuous variables. The categorical variables were described as frequencies and percentages, while continuous variables were presented as medians and interquartile range (IQR) values and means and standard deviations. Means between groups were compared using One-way analysis of variance (ANOVA). A Kruskal-Wallis H test was used to compare the medians of the different group laboratory parameters. Categorical variables were analyzed by using the chi-square test, and wherever the data were limited, Fisher Exact test was used. Differences between groups means and medians are considered significant when a p value is < 0.05. All statistical analyses were performed using GraphPad Prism software (La Jolla, CA, USA) and SPSS (Statistical Package for Social Sciences) for Windows version 25.0 (IBM SPSS Inc., USA).

## Data Availability

Data available on request from the authors

## Acknowledgment

We would like to thank the adminstration of Jaber Al-Ahmad Hospital for their help and support.

## Author Contribution

AA1: Data collection and analysis, critical revision, HA: Study conception and design, data analysis and manuscript writing, YA: Data collection, TA: Data collection, MA: Data analysis, JA: Data analysis, SD: data analysis, AAD: Data analysis, AB: Data collection from ICU, HT: Data collection for pediatric patients, AA2: Data collection, NA: Data collection, EA: Data analysis, FA: Data analysis, critical revision, MB: Data collection

## References

1 Huang, C. et al. Clinical features of patients infected with 2019 novel coronavirus in Wuhan, China. Lancet 395, 497–506, doi:10.1016/S0140-6736(20)30183-5 (2020).

2 Wu, F. et al. A new coronavirus associated with human respiratory disease in China. Nature 579, 265–269, doi:10.1038/s41586-020-2008-3 (2020).

3 Dong, E., Du, H. & Gardner, L. An interactive web-based dashboard to track COVID-19 in real time. Lancet Infect Dis, doi:10.1016/S1473-3099(20)30120-1 (2020).

4 Bai, Y. et al. Presumed Asymptomatic Carrier Transmission of COVID-19. JAMA, doi:10.1001/jama.2020.2565 (2020).

5 Lai, C. C. et al. Asymptomatic carrier state, acute respiratory disease, and pneumonia due to severe acute respiratory syndrome coronavirus 2 (SARS-CoV-2): Facts and myths. J Microbiol Immunol Infect, doi:10.1016/j.jmii.2020.02.012 (2020).

6 Wang, D. et al. Clinical Characteristics of 138 Hospitalized Patients With 2019 Novel Coronavirus-Infected Pneumonia in Wuhan, China. JAMA, doi:10.1001/jama.2020.1585 (2020).

7 Lu, X. et al. SARS-CoV-2 Infection in Children. N Engl J Med 382, 1663–1665, doi:10.1056/NEJMc2005073 (2020).

8 Salje, H. et al. Estimating the burden of SARS-CoV-2 in France. Science, doi:10.1126/science.abc3517 (2020).

9 Al-Mulla, F. et al. A comprehensive germline variant and expression analyses of <em>ACE2</em>, <em>TMPRSS2</em> and SARS-CoV-2 activator <em>FURIN</em> genes from the Middle East: Combating SARS-CoV-2 with precision medicine. bioRxiv, 2020.2005.2016.099176, doi:10.1101/2020.05.16.099176 (2020).

10 Benetti, E. et al. ACE2 gene variants may underlie interindividual variability and susceptibility to COVID-19 in the Italian population. medRxiv, 2020.2004.2003.20047977, doi:10.1101/2020.04.03.20047977 (2020).

11 Hussain, M. et al. Structural variations in human ACE2 may influence its binding with SARS-CoV-2 spike protein. J Med Virol, doi:10.1002/jmv.25832 (2020).

12 Devaux, C. A., Rolain, J. M. & Raoult, D. ACE2 receptor polymorphism: Susceptibility to SARS-CoV-2, hypertension, multi-organ failure, and COVID-19 disease outcome. J Microbiol Immunol Infect 53, 425–435, doi:10.1016/j.jmii.2020.04.015 (2020).

13 Al-Shammari, A. A. A. et al. Real-time tracking and forecasting of the COVID-19 outbreak in Kuwait: a mathematical modeling study. medRxiv, 2020.2005.2003.20089771, doi:10.1101/2020.05.03.20089771 (2020).

14 Organization, W. H. (World Health Organization, 2020).

15 Force, A. D. T. et al. Acute respiratory distress syndrome: the Berlin Definition. JAMA 307, 2526–2533, doi:10.1001/jama.2012.5669 (2012).

16 Levey, A. S. et al. Definition and classification of chronic kidney disease: a position statement from Kidney Disease: Improving Global Outcomes (KDIGO). Kidney Int 67, 2089–2100, doi:10.1111/j.1523-1755.2005.00365.x (2005).

17 Guan, W. J. et al. Clinical Characteristics of Coronavirus Disease 2019 in China. N Engl J Med 382, 1708–1720, doi:10.1056/NEJMoa2002032 (2020).

18 Bureau, K. C. S. Population estimates in Kuwait by Age, Nationality and sex at 1L1L2019. (2019).

19 Byambasuren, O. et al. Estimating the extent of asymptomatic COVID-19 and its potential for community transmission: systematic review and meta-analysis. medRxiv, 2020.2005.2010.20097543, doi:10.1101/2020.05.10.20097543 (2020).

20 Yang, R., Gui, X. & Xiong, Y. Comparison of Clinical Characteristics of Patients with Asymptomatic vs Symptomatic Coronavirus Disease 2019 in Wuhan, China. JAMA Netw Open 3, e2010182, doi:10.1001/jamanetworkopen.2020.10182 (2020).

21 Gandhi, M., Yokoe, D. S. & Havlir, D. V. Asymptomatic Transmission, the Achilles’ Heel of Current Strategies to Control Covid-19. N Engl J Med 382, 2158–2160, doi:10.1056/NEJMe2009758 (2020).

22 Spychalski, P., Blazynska-Spychalska, A. & Kobiela, J. Estimating case fatality rates of COVID-19. Lancet Infect Dis, doi:10.1016/S1473-3099(20)30246-2 (2020).

23 Richardson, S. et al. Presenting Characteristics, Comorbidities, and Outcomes Among 5700 Patients Hospitalized With COVID-19 in the New York City Area. JAMA, doi:10.1001/jama.2020.6775 (2020).

24 Jin, J. M. et al. Gender Differences in Patients With COVID-19: Focus on Severity and Mortality. Front Public Health 8, 152, doi:10.3389/fpubh.2020.00152 (2020).

25 Rahimi-Levene, N. et al. Lower hemoglobin transfusion trigger is associated with higher mortality in patients hospitalized with pneumonia. Medicine (Baltimore) 97, e0192, doi:10.1097/MD.0000000000010192 (2018).

26 Arachchillage, D. R. J. & Laffan, M. Abnormal coagulation parameters are associated with poor prognosis in patients with novel coronavirus pneumonia. J Thromb Haemost 18, 1233–1234, doi:10.1111/jth.14820 (2020).

27 Bowles, L. et al. Lupus Anticoagulant and Abnormal Coagulation Tests in Patients with Covid-19. N Engl J Med, doi:10.1056/NEJMc2013656 (2020).

28 Garcia-Olive, I., Sintes, H., Radua, J., Abad Capa, J. & Rosell, A. D-dimer in patients infected with COVID-19 and suspected pulmonary embolism. Respir Med 169, 106023, doi:10.1016/j.rmed.2020.106023 (2020).

29 Lippi, G. & Plebani, M. Procalcitonin in patients with severe coronavirus disease 2019 (COVID-19): A meta-analysis. Clin Chim Acta 505, 190–191, doi:10.1016/j.cca.2020.03.004 (2020).

30 Li, X. et al. Clinical characteristics of 25 death cases with COVID-19: A retrospective review of medical records in a single medical center, Wuhan, China. Int J Infect Dis 94, 128–132, doi:10.1016/j.ijid.2020.03.053 (2020).

31 Soler, M. J., Wysocki, J. & Batlle, D. ACE2 alterations in kidney disease. Nephrol Dial Transplant 28, 2687–2697, doi:10.1093/ndt/gft320 (2013).

32 Ronco, C., Reis, T. & Husain-Syed, F. Management of acute kidney injury in patients with COVID-19. Lancet Respir Med, doi:10.1016/S2213-2600(20)30229-0 (2020).

33 Awad, A. & Al-Nafisi, H. Public knowledge of cardiovascular disease and its risk factors in Kuwait: a cross-sectional survey. BMC Public Health 14, 1131, doi:10.1186/1471-2458-14-1131 (2014).

34 Mennuni, S. et al. Hypertension and kidneys: unraveling complex molecular mechanisms underlying hypertensive renal damage. J Hum Hypertens 28, 74–79, doi:10.1038/jhh.2013.55 (2014).

